# Self-experience in MDMA assisted therapy of PTSD

**DOI:** 10.1101/2023.01.03.23284143

**Authors:** Bessel A. van der Kolk, Julie B. Wang, Rachel Yehuda, Leah Bedrosian, Allison Cooker, Charlotte Harrison, Michael Mithoefer, Berra Yazar-Klosinki, Amy Emerson, Rick Doblin

**Affiliations:** Trauma Research Foundation, Brookline, MA, USA; MAPS Public Benefit Corporation (MAPS PBC), San Jose, CA, USA; James J. Peters Veterans Affairs Medical Center, Bronx, NY USA; Center for Psychedelic Psychotherapy and Trauma Research, Icahn School of Medicine at Mount Sinai, New York, NY, USA; Department of Psychiatry and Behavioral Sciences, Medical University of South Carolina, Charleston, SC, USA; University of California, San Francisco, San Francisco, CA, USA; Multidisciplinary Association for Psychedelic Studies (MAPS), San Jose, CA, USA

## Abstract

In recent years there has been a resurgence of interest in the therapeutic potential of psychedelic substances such as 3,4-methylenedioxymethamphetamine (MDMA). This renaissance of psychedelic studies opens the door for a new paradigm in psychiatric medicine: drug-facilitated psychotherapy. In this study we report the findings of a randomized, double-blind, placebo-controlled, multi-site Phase 3 clinical trial (NCT03537014) to test the effects of MDMA-assisted therapy (MDMA-AT) on patients with severe PTSD. The vast majority (85%) of individuals in this study reported having suffered early childhood trauma, which is strongly associated with deficits in emotional coping skills /altered self-capacities, which have been shown to constitute major obstacles to successful completion of currently available evidence-based treatments. Partcicipants were randomized 1:1 to receive manualized therapy with either MDMA or placebo with three preparatory and nine integrative therapy sessions. Symptoms were measured at baseline and at 2□ months after the last experimental session with the Clinician-Administered PTSD Scale for DSM-5, the Toronto Alexithymia Scale (TAS_20), the Self Compassion Scale (SCS) and the Inventory of Altered Self-Capacities (IASC). MDMA-AT, compared with psychotherapy alone, significantly altered the domains of alexithymia, self-compassion, and altered self-capacities. These findings suggest that MDMA-AT can substantially improve transdiagnostic mental processes associated with poor treatment response.

## Introduction

In recent years there has been a resurgence of interest in the therapeutic potential of psychedelic substances such as tryptamines (e.g., psilocybin), ketamine and phenethylamines (e.g., 3,4-methylenedioxymethamphetamine (MDMA)^1, 2^,. The renaissance of psychedelic studies opens the door for a new paradigm in psychiatric medicine: drug-facilitated psychotherapy.

A pooled analysis of six MDMA-assisted therapy Phase II trials showed that 54% of patients no longer met criteria for PTSD^3^. Based on its positive performance with significant and sustained reductions in PTSD symptoms and acceptable safety profiles the FDA has designated MDMA-assisted therapy (MDMA-AT) as a breakthrough therapy for PTSD^4^. Recently, results of a Phase 3 multisite study of MDMA assisted psychotherapy were published confirming the safety and efficacy of MDMA-AT in individuals with severe PTSD^5^. Compared with the placebo with therapy (P+Th) condition MDMA-AT was found to induce significant and robust attenuation in PTSD symptom severity score (P<□ 0.0001, d=□ 0.91), suggesting a strikingly greater therapeutic effect of MDMA-AT over protocolized therapy alone.

The protocol for MDMA-assisted therapy consists of a 3-month long treatment with 3 dosing and 3 preparation sessions, as well as 9 integration sessions. All study participants received an equal, substantial dose of manualized therapy in addition to receiving either the MDMA or placebo. This provides us with an opportunity to explore the differential effects of therapy alone vs psychedelic-assisted therapy to gain a deeper understanding of the psychological change processes induced by psychedelic therapies.

Trauma-focused psychotherapy is considered a first line treatment for PTSD^6^,^7^. However, the overall success rate with psychotherapeutic treatments for PTSD has been relatively disappointing. At least one-quarter of patients drop out of trauma-focused psychotherapy, and up to one-half are left with significant lingering symptoms ^8^,^9, 10^. Even patients who are considered responders often remain challenged by difficulties in emotion regulation, impulse control and interpersonal functioning^11^, ^12, 13^, all of which seem to continue relatively independent from PTSD symptomatology^14, 15^.

Many trauma survivors, particularly those with histories of child abuse (developmental trauma) have been shown to experience significant defects in a variety of transdiagnostic mental processes, including a loss of sense of safety, trust and self-worth, being unable to notice internal states (alexithymia), lack of a coherent sense of self, inability to modulate or tolerate distress, difficulties negotiating interpersonal conflicts and negative self-appraisals, such as shame, self-blame and lack self-compassion. All of these have been shown to correlate with poor treatment outcome^16, 17^.

Multiple studies have shown that reduced self-capacities interfere with successful completion of psychotherapy for PTSD^18^,^19^. Problems with emotion regulation interfere with being able to disengage from trauma-related stimuli, which increases the probability of drop out due to an inability to manage distress arising during treatment ^20^. Alexithymia, deficits in being able to identify and describe emotions, is associated with posttraumatic pathology^21, 22, 23^ and with a lack of habituation to emotionally distressing stimuli^24^. Persons with high alexithymia scores have been shown to display low autonomic activity in response to any task performance, regardless of the level of emotional demand, including processing traumatic maternal^25^.

Finally, self-compassion is a core component of overall mental health and well-being^26^ often lacking in trauma survivors with PTSD who frequently experience self-loathing and self-blame^27, 28^. Low self-compassion scores are associated with anxiety, depression, narcissism, self-criticism, and with poor treatment responses^29^. Stabilizing self-experience that leads to higher levels of emotion-regulation and self-compassion has been shown to improve treatment results for a variety of psychological interventions^30, 31^.

In this paper, we report the results of three transdiagnostic outcome measures that were collected in tandem with the previously published PTSD changes in the MDMA-AT Phase 3 trial. This provides us with an opportunity to illuminate psychological processes that underpin the significant gains and sustained effects of MDMA-AT compared with therapy alone.

## Methods

### Study Design

This paper assesses exploratory data from a randomized, double-blind, placebo-controlled study comparing safety and efficacy of MDMA-AT to inactive placebo with therapy (P+TH) in participants with severe PTSD^5^. Details such as recruitment and locations of the 15 sites are described in the previous paper. All participants, site staff, independent raters, and the sponsor were blind to participants group assignments until after database lock. All participants provided written informed consent at eligibility screening after ethics approval from local Institutional Review Boards.

### Participants

All 90 participants met DSM-5 criteria for current PTSD with a symptom duration of six months or greater and a CAPS-5 total severity score of 35 or greater at baseline. The vast majority of participants (85%) suffered from developmental trauma (childhood physical and/or sexual abuse) and 87% had experienced multiple traumas. Exclusion criteria included primary psychotic, bipolar I, dissociative identity, personality disorders, current alcohol and substance use disorders, and any medical condition for which an acute, transient increase in blood pressure or heart rate would pose a medical concern. Full eligibility criteria are described in the study protocol (http://maps.org/mapp1).

### Intervention

Participants underwent three 90-minute preparatory therapy sessions with a co-therapist dyad to establish therapeutic alliance and prepare for experimental sessions. The treatment period consisted of three 8-hour experimental sessions of either MDMA-AT or inactive placebo control with the same therapy, spaced approximately four weeks apart, also described previously^6^. In each experimental session, participants were given a divided-dose of MDMA or placebo, with an initial dose followed by a supplemental half-dose 1.5 to 2.5 hours later. In the first experimental session the dose was 80 mg + 40 mg MDMA HCl, and in second and third experimental sessions, the dose was escalated to 120 mg + 60 mg MDMA HCl. Manualized therapy was conducted in accordance with MAPS MDMA-AT treatment manual (maps.org/treatment manual). Following each experimental session, participants underwent three 90-minute integration sessions, scheduled one week apart, to provide them with the opportunity to process their experiences.

### Demographic and Baseline Variables

Age, gender, ethnicity, race, and education were compared between treatment groups. Other variables relevant to the transdiagnostic outcomes explored here, but not reported in this publication, included employment status, detailed trauma history, pre-study treatment, and baseline outcomes measures for the Adverse Childhood Experience Questionnaire (ACE)^32^, Beck Depression Inventory II (BDI-II)^33^, Clinician-Administered PTSD Scale for DSM-5 (CAPS-5) total severity score^34^, and lifetime suicidality assessment from the Columbia Suicide Severity Rating Scale (C-SSRS) ^35^.

### Self-experence Measures

#### The Inventory of Altered Self Capacities (IASC)

a 63-item self-report measure of difficulties with relationships, identity, and regulation, rated on a 5-point Likert scale ranging from 1 (“Never”) to 5 (“Very Often”). The IASC consists of the following subscales: Interpersonal Conflicts, Identity Impairment, Idealization Disillusionment, Abandonment Concerns, Susceptibility to Influence,

Affect Dysregulation (with two subscales: Affect Skill Deficits and Affect Instability), and Tension Reduction Activities. Items for each subscale are summed to calculate subscale raw scores that range from 9 to 45^36^.

#### The Toronto Alexithymia Scale (TAS-20)

a 20-item measure of self-reported difficulties with recognizing and verbalizing emotions. Responses are reported on a 5-point Likert scale ranging from 1 (“Strongly disagree”) to 5 (“Strongly agree”). The scale is comprised of three subscales: Difficulty Describing Feelings,

Difficulty Identifying Feelings, and Externally-Oriented Thinking. Total scores diagnostically indicate no alexithymia (≥50), border-line alexithymia (51-60), and alexithymia (≥61)^37^.

#### The Self-Compassion Scale (SCS)

a 26-item self-report measure of how respondents perceive their own failures, suffering, or inadequacies with kindness and compassion as a part of the common human experience. Respondents indicate how they often feel for each item on a 5-point Likert scale ranging from 1 (“Almost never”) to 5 (“Almost always”). The SCS consists of six subscales: Self-Kindness, Self-Judgement, Common Humanity, Isolation, Mindfulness, and Over-Identified. The mean of subscale scores serves as the total score^38^. A total score of 1-2.4 indicates “low,” 2.5-3.4 “moderate,” and 3.5-5.0 “high” SCS.

Independent raters conducted the PTSD primary outcome assessment, CAPS-5, prior to the first experimental session and at the primary endpoint Visit 19, approximately eight weeks after the final experimental session (18 weeks post-baseline). The SCS, IASC, and TAS-20 were self-reported at baseline, during the final preparatory session (Visit 4) and again approximately 18 weeks later at study termination (Visit 20).

### Statistical Methods

Descriptive analyses were performed on demographic, baseline, and outcome variables. Group means (SD) were compared using *t*-tests or ANOVA/ ANCOVA and proportions were compared using chi-square tests. Non-parametric tests were performed on samples with non-normal distributions. Pearson’s correlations were conducted to examine linear relationships across variables.

In the primary analysis, separate two-way ANCOVA models, adjusting for baseline scores and CAPS-5 dissociative subtype (Yes=1 and No=0), compared treatment group differences in change scores for TAS-20, SCS, each IASC factor, and CAPS-5 (MDMA-AT vs. P+Th).

Separate analyses examined within-subjects differences at baseline and follow-up scores for MDMA-AT and P+Th groups. Sub-set analyses evaluated change scores stratified by baseline cutoff scores; specifically: (i) TAS-20 baseline measure of having no alexithymia (≤50) and alexithymia (>51); (ii) SCS baseline measure of low (1-2.4) and moderate (2.5-3.4) or high (3.5-5.0) self-compassion; (iii) and for each IASC factor baseline scores for each factor above and below the sample median For IASC factors, the sample median (vs. the mean) was used to account for any non-normal sample distributions and since the IASC lacks a validated composite score to define a clinical cutoff. Models tested the main effects and interaction terms between treatment group (MDMA-AT vs. P+Th) and baseline categories (low vs. high TAS-20, SCS, or IASC factor). All models adjusted for baseline scores and CAPS-5 dissociative subtype (Yes=1 and No=0). Tukey’s HSD test corrected for multiple comparisons and tables reported Least Square Means (LSMEANS) which adjusted for unequal sample sizes across group comparisons. All analyses were performed using SAS Version 9.4 (SAS Institute, Cary, North Carolina).

## Results

### Sample Characteristics

The study sample consisted of 90 participants who were randomized and completed at least one experimental dosing session (MDMA=46, P+Th=44). Follow-up data for TAS-20, SCS, and IASC were missing for eight participants due to early study termination (discontinued due to COVID-19 = 4; declined further treatment = 2; restarted pre-study treatment = 1). All available data were used in the analysis. Participants were majority women (63.3%), White (78.4%), non-Hispanic or Latino (90.0%), college graduates (71.1%), and the mean (SD) age was 40.93 (11.95) years. 85% of participants had histories of childhood physical/sexual abuse (developmental trauma), and 87% had suffered multiple traumas. There were no statistically significant group differences between MDMA-AT and P+Th groups across demographic and baseline variables. Detailed sample characteristics have been described in the primary outcome paper^7^

### Treatment Effects on Self-experience Measures

Most study participants had significant improvements in the measures of self-experience. Those with high baseline levels of alexithymia, low self-compassion, and/ or altered self-capacities had significant improvement at follow-up; and these improvements were more pronounced in the MDMA-AT (vs. P+Th) group **(Table 1)**. MDMA-AT had a greater improvement than P+Th on alexithymia [-12.85 (2.43) vs. -1.51 (2.26); *p* < .0006] **(figure 1)**, self-compassion 1.36 (0.20) vs. 0.23 (0.16); *p* < .0001] (**figure 2**), and IASC factors: “idealization disillusionment” [-0.75 (0.21) vs. -0.20 (0.17); *p* = .05], “susceptibility to influence” [-0.56 (0.15) vs. 0.02 (0.15); *p* = .0099], and “affect skill deficit” [-1.60 (0.92) vs. - 0.84 (1.45); *p* = .02] (**figure 3**). This suggests that MDMA has a strong effect of these measures of emotion regulation and self-experience, even after adjusting for potential covariates and conducting multiple comparisons.

**Table 1:**
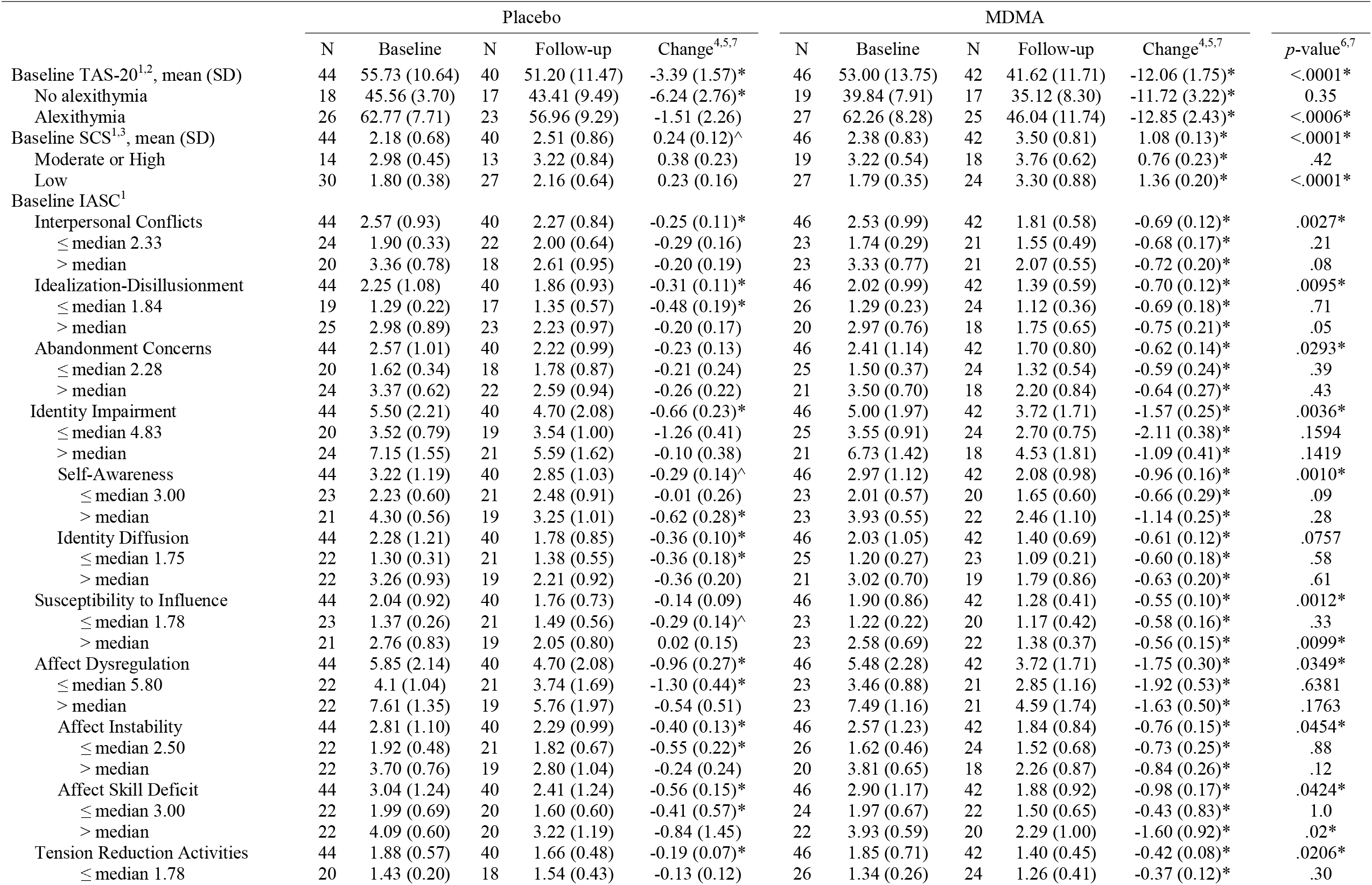
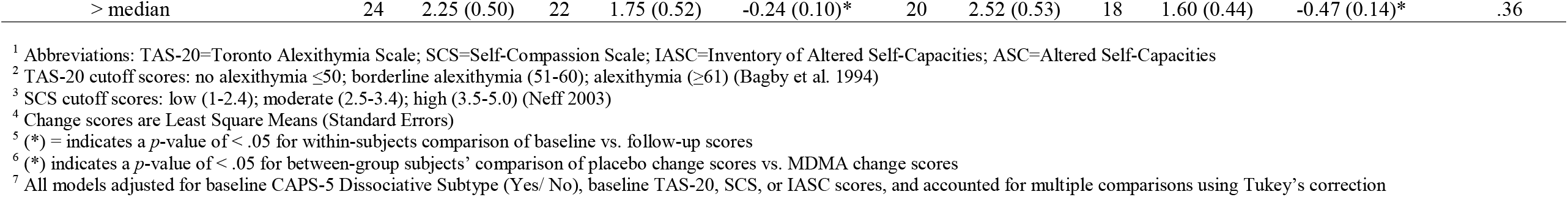
Mean (SD) or LSM (SE) for Alexithymia (TAS-20), Self-Compassion (SCS), and Inventory of Altered Self-Capacities (IASC) Scores by Low/ High Baseline Scores.

**Fig 1.**
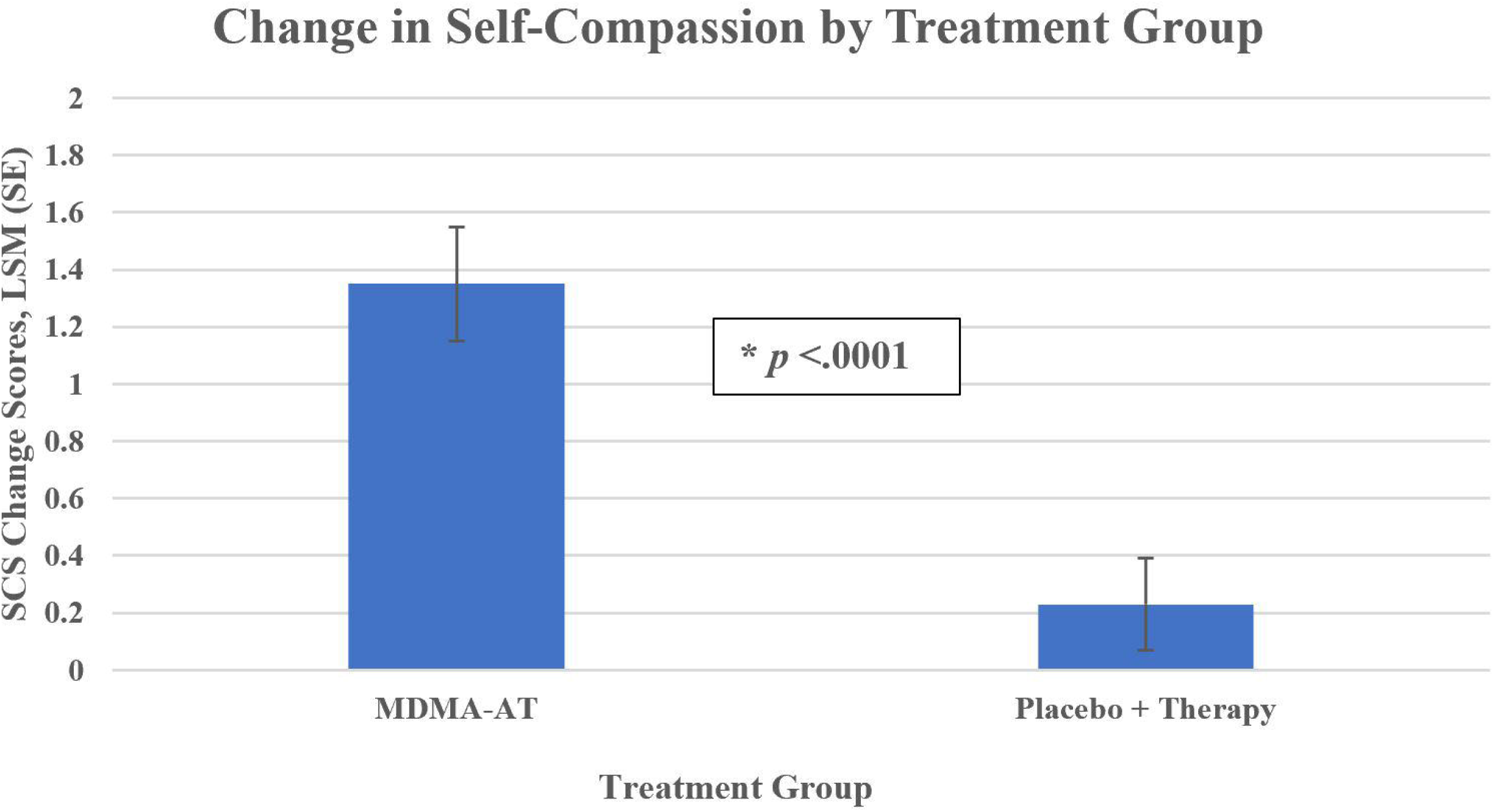
Alexithymia Change scores in MDMA-AT. Least square means ((SE) chmage in Toronto Alexithymkia Scale (TAS-20) nscore4s from baseline to follow up by treatment group: MDMA-AT=.-12.85 (2.43) vs P+Th+-1.51 (2.26), p<.0001.

**Fig 2.**
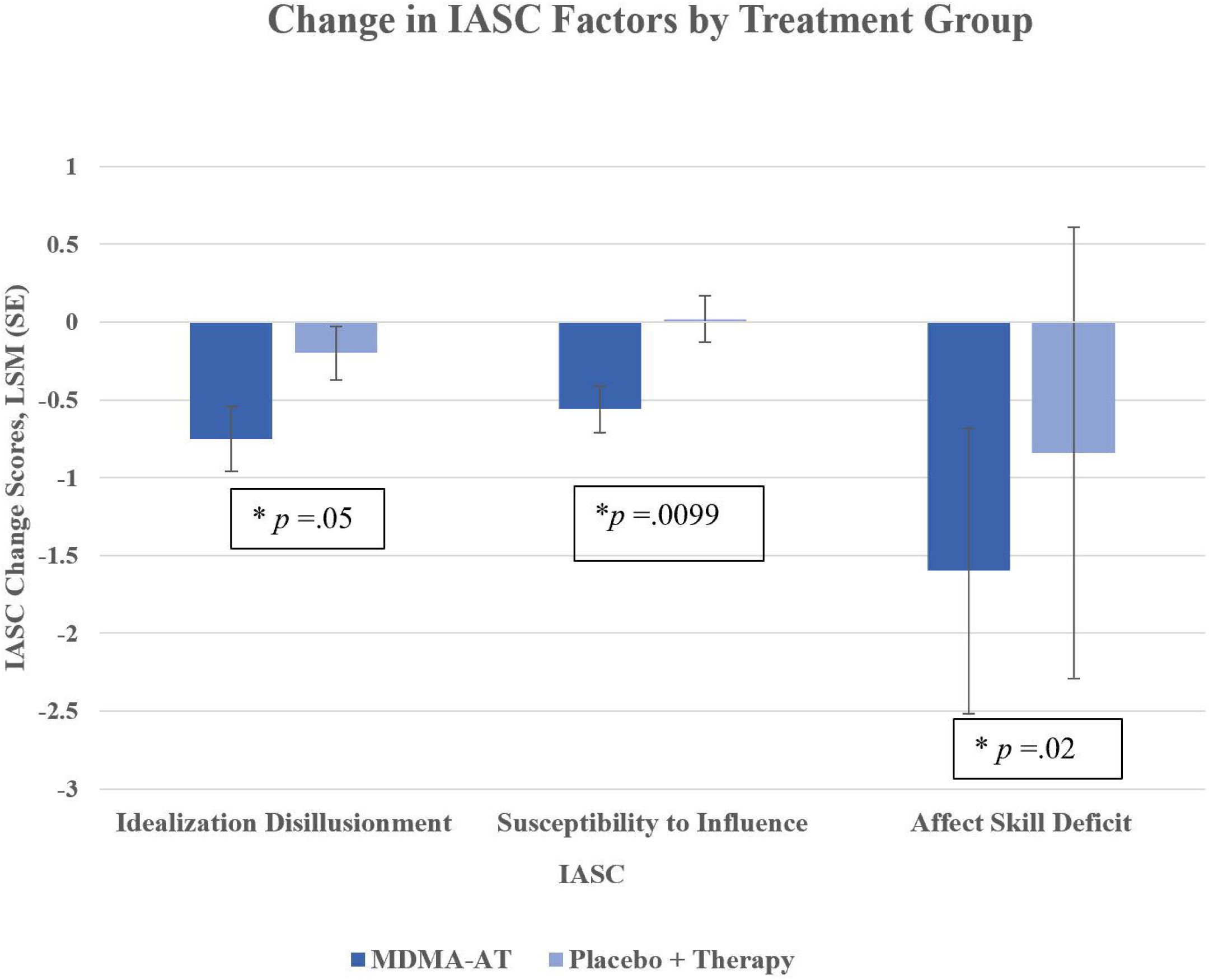
Self-compassion Chgange Scores in MDMA-AT. Least square means (SE) change in Self-compassion Scale (SCS) from baseline to foillow-up bby treatment group: MDMA-AT= 1.35(0.20) vs. P+Th=0.23 (0.16), P<.0001.

**Fig 3.**
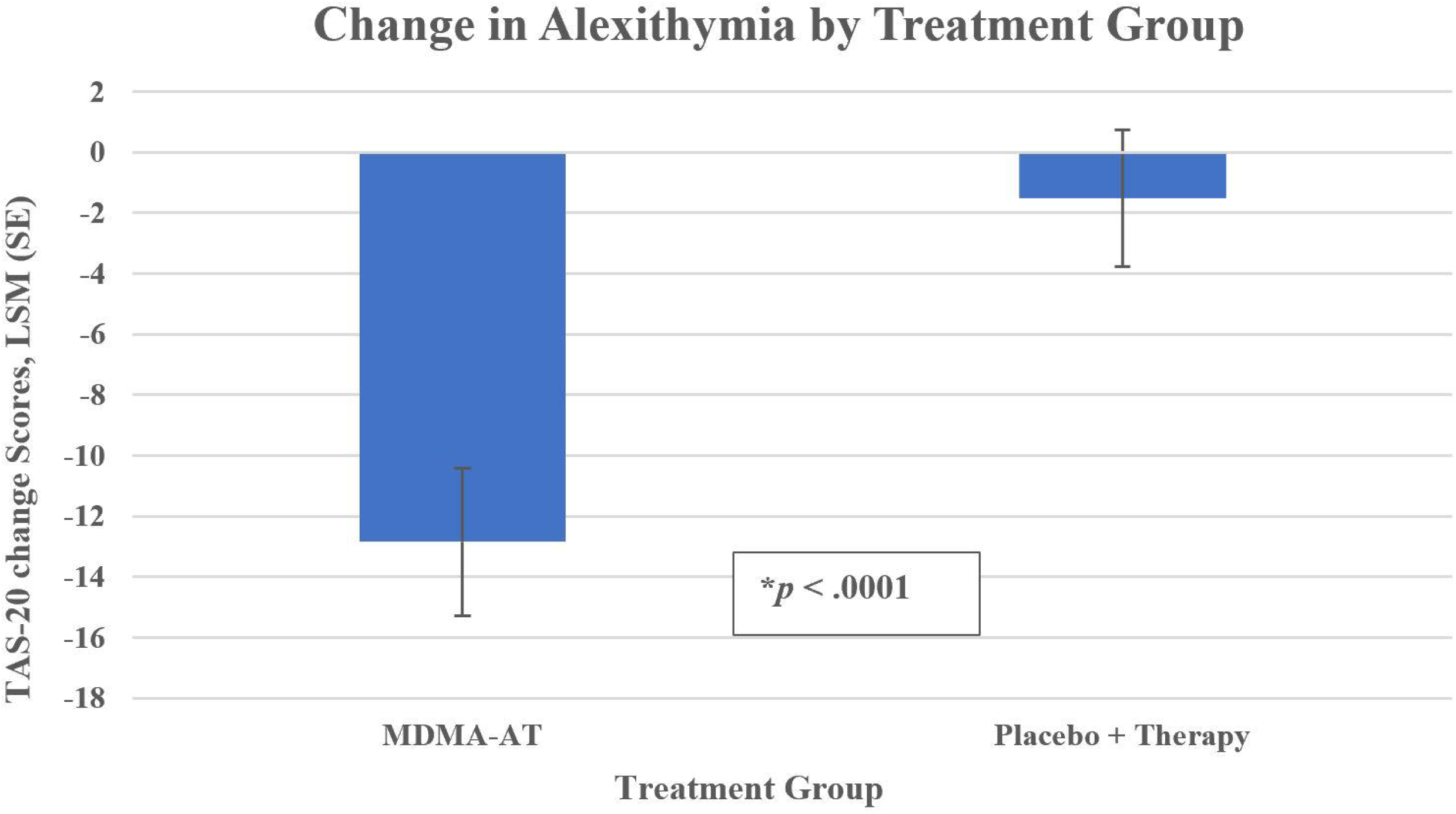
Inventory of Altered Self-capacities (IASC) Change Scores. Least square means (SE) change from baseline to follow-up by treatment group: (a) ‘Idealization Disillusionment’ MDMA-AT = -0.75 (0.21) vs. Placebo+Th = -0.20 (0.17), *p* = .05; (b) ‘Susceptibility to Influence’ MDMA-AT = -0.56 (0.15) vs. Placebo+Th = 0.02 (0.15), *p* = .0099; and (c) ‘Affect Skill Deficit’ MDMA-AT = -1.60 (0.92) vs. Placebo+Th = -0.84 (1.45), *p* = .02.

### Baseline self-capacity Measures & Treatment Effects on PTSD Symptoms

Several baseline measures of self-experience predicted CAPS-5 change scores in the MDMA-AT group: those worse off at baseline had greater treatment effects than the placebo. Group (Table 2). There was a greater reduction in CAPS-5 scores in the MDMA-AT group for those who had begun the trial with greater baseline alexithymia (−16.16; 95% CI: -28.80, -7.52), and poorer SCS scores (−13.85; 95% CI:-22.84, -4.86*)., Participants who started with higher self-capacities on the IASC benefited more from MDMA, specifically in “idealization disillusionment” [-12.80; 95% CI: -22.61, -2.98], “identity impairment” [-11.87; 95% CI: -21.18, -2.56], “identity diffusion” [-12.56; 95% CI: -21.92, -3.21], and “susceptibility to influence” [-13.96; 95% CI: -23.68, -4.24].

**Table 2:**
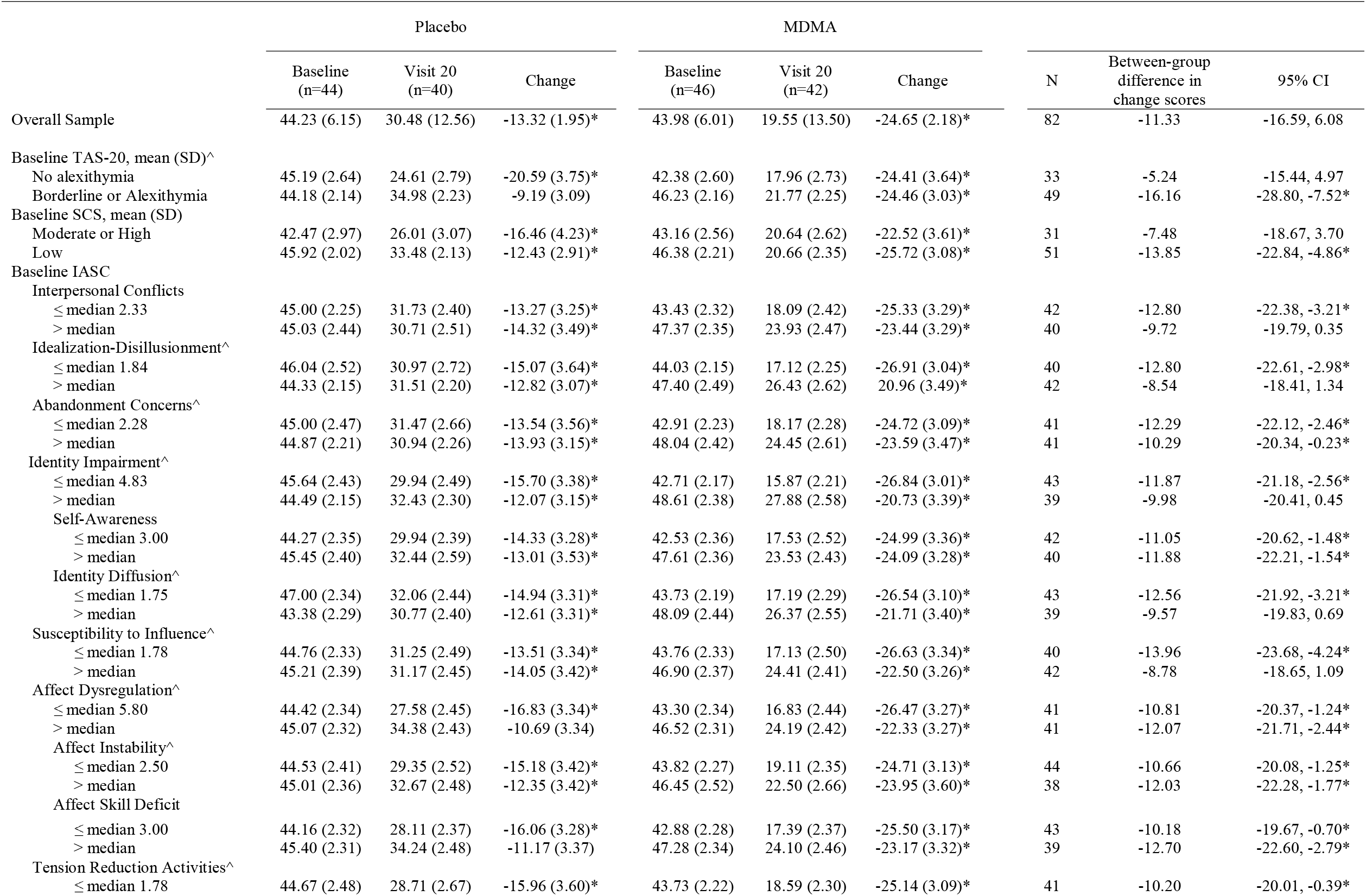
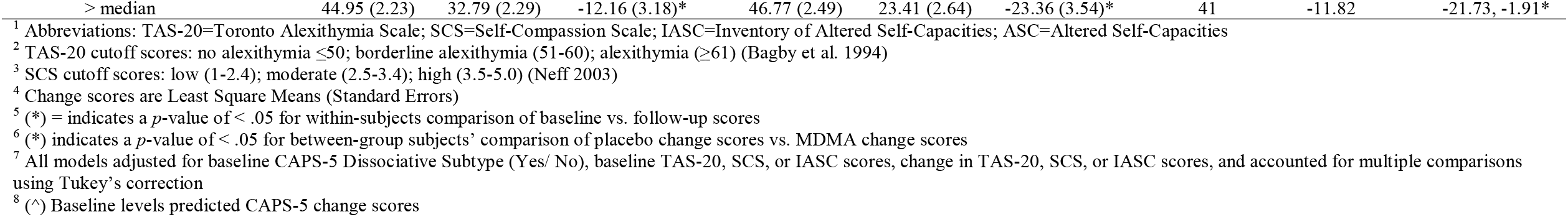
LSM (SE) CAPS-5 Total Severity Scores, Change Scores, and Difference in Change Scores by Low/ High TAS-20, SCS, and IASC Baseline Scores.

## Discussion

This study of 90 participants with chronic PTSD demonstrated that, in addition to improving PTSD symptmatology, administration of MDMA in conjunction with therapy also significantly improved measures of self-experience, including affect dysregulation, negotiation of interpersonal conflicts, alexithymia and self-compassion, when compared to treatment with therapy plus placebo. The change scores in measures of self-experience were highly corelated with recovery from PTSD (e.g. the TAS-20 total change scores predicted CAPS-5 total severity change scores at the p < .0001 level). Endpoint emotion regulation and self-capacities were critical factors predicting remission of PTSD. Treatment responders in both the MDMA-AT and the P+Th condition had a statistically significant difference in TAS-20, SCS and IASC change scores from CAPS-5 non-responders, confirming the notion that emotion regulation and self-capacities are critical elements in positive treatment outcome.

While both treatment conditions were associated with some improvement in self-experience, only MDMA-AT, and not psychotherapy alone, significantly altered these transdiagnostic measures, with significantly greater changes in the domains of alexithymia, self-compassion, emotion regulation, negotiating interpersonal conflicts, abandonment concerns, self-awareness, idealization/disillusionment, susceptibility to influence, and tension reduction activities.

The clinical relevance of these self-experience measures for treatment outcome is illustrated by the finding that among participants in the therapy/placebo group only those who started with adequate scores on the various self-experience scales had a significant improvement in their PTSD scores. In contrast, in participants with low self-capacity ratings at baseline only the MDMA-AT condition produced significant improvements in PTSD, in tandem with significant change scores in TAS-20, and SCS ratings, and in 8 of 9 IASC factors change scores, including interpersonal conflicts, lack of self-awareness and tension reduction activities (see Table 3 in the Appendix). Thus, MDMA-AT appears to substantially improve mental processes associated with resilience and positive response to treatment.

MDMA has been shown to promote a general sense of interpersonal “connectedness” ^39^, and “openness” ^40^, and to enhance positive appraisal of favorable memories, while reducing negative evaluations of painful memories^41^. It also has been shown to enhance extinction of fearful memories, modulate memory reconsolidation (possibly through an oxytocin-dependent mechanism), and to promote social behavior ^42^. Moreover, MDMA inhibits habitual fear responses to emotional threats^43^. This is thought to facilitate being able to put the emotional sequelae of painful past experiences into a realistic perspective.

Most of the studies of mental changes secondary to the administration of MDMA have been conducted in normal populations who are less likely to suffer from major problems with self-experience. In this study, we examined the effects of MDMA on a group of individuals with major clinical deficits in domains that have previously been investigated mainly in non-clinical populations and that have been found to be associated with treatment resistance. Our findings suggest that the therapeutic benefits of MDMA may be most pertinent for persons with clinically significant impairment in emotion regulation and self-capacities.

The vast majority (85%) of traumatized individuals in this study reported having suffered early childhood trauma, i.e. physical or sexual abuse by their caregivers. Only 4 out of 90 subjects in this study had an Adverse Childhood Experience (ACE) score of 0. Histories of child maltreatment are associated with poorer responses to psychotherapy in individuals diagnosed with PTSD^44, 45^. Abuse at the hands of one’s early caregivers has been shown to put individuals at risk for deficits in emotional coping skills /altered self-capacities, major obstacles to successful completion of currently available evidence-based treatments^46, 47^.

Being able to emotionally process traumatic experiences is an important element of successful treatment^48, 49^. Being able to identify feelings, describing them and recognizing their triggers allows an individual to reflect on the situation and to respond appropriately to the context, rather than acting solely on their emotional arousal^50^. For example, alexithymia, avoidance of distressing wishes, feelings or experiences, and trouble recalling distressing experiences, is associated with impaired affect regulation^22, 23, 24^.

Alexithymia has frequently been observed in the context of invalidating or abusive early environments where children learn that communicating emotional experiences is inappropriate, ineffective, or potentially dangerous^51, 52^. Unable to escape physically from chronic abuse, alexithymic individuals are thought to have learned to disengage from both their external reality as well as their internal experiences^53^.

Even though the day-long MDMA-AT sessions often occurred in relative silence as participants focus largely on their inner experience, MDMA was associated with a significant improvement in emotional self-awareness and loss of alexithymia. This suggests that MDMA can facilitate accessing painful memories and experiences that under ordinary conditions are too overwhelming and terrifying to confront.

Adaptive emotion regulation is essential for effective treatment of PTSD. Trauma-focused treatments for PTSD require both activation and modification of fearful memories. This activation depends on two processes: physiological reactivity to trauma-related stimuli, and being able to tolerate the subjective distress generated by these traumatic memories^54^. Being able to tolerate physiological arousal to trauma-related stimuli predicts improvement in exposure treatment, supporting a gradual diminution in the distress experienced in response to trauma recall (habituation) within- and between-sessions^55^.

Emotion regulation (ER) deficits are major contributors to the development of a large variety of psychopathological conditions^56^, including interference with being able to resolve the impact of traumatizing experience(s) ^57^,^58, 59^,^60^. Whereas healthy, flexible ER capacities are key factors underlying well-being, ER difficulties comprise a transdiagnostic risk factor for mental health problems in general, including the development and/or maintenance of symptoms of PTSD ^61^, by interfering with being able to disengage from trauma-related stimuli and inhibiting maladaptive emotion regulation strategies^62^. Problems with emotion regulation influence both the development and the maintenance of PTSD symptoms after exposure to potentially traumatizing experiences, ^63, 64, 65^, and predict both functional impairment and symptom complexity^66^.

Self-compassion is another core component of overall mental health and well-being^26^. Individuals suffering from traumatic stress often suffer from shame, self-blame and self-loathing^28, 29^. Appraisals of mental defeat and permanent change have a profound and debilitating effect on an individual’s identity and sense of self ^67^. Low self compassion scores have consistently been associated with symptoms such as anxiety, depression, narcissism, self criticism and avoidance^26, 29, 59^. Being caring and kind to oneself, rather than critical, even under stress, can mitigate the negative effects of trauma exposure by increasing resilience and by decreasing avoidance-oriented coping ^68, 69^. Self-compassion has been shown to boost the efficacy of cognitive reappraisals^70^.

### Summary

Defective self-capacities seem to be major obstacles to successful completion of currently available evidence-based treatments of PTSD, making the development of innovative treatments that address those capacities a research priority. This study suggests that MDMA may be particularly effective for enhancing treatment efficacy by improving a range of problems with self -experience that are associated with treatment resistance. Assessment of self-capacities may be as relevant for treatment planning and outcome research as measuring PTSD severity, because, as this study suggests, psychotherapy alone may not sufficiently compensate for the debilitating effects of deficient self-experience on being able to confront traumatic material and thus, on treatment outcome.

#### Limitations

This study sample was not focused on treatment resistant individuals and did not control for age and nature of trauma exposure The striking lack of correlation between baseline ACE and TAS-20, SCS, and IASC factors likely is due to lack of variability in the ACE measurement scale that ranged from 1-10 where the sample mean (SD) was 5.0 (2.8). In this study only 4 participants indicated an ACE score of 0. In this Phase 3 MDMA-AT trial there has not yet been a long term follow-up of the sustainability of treatment gains in this population More studies are needed to examine the capacity of MDMA to ameliorate post-traumatic symptomatology in a variety of trauma populations, including whether MDMA treatment is capable of permanently altering a host of psychological processes associated with having been traumatized, including shame, self-blame, the capacity for emotional intimacy, executive functioning and affect regulation.

## Data Availability

All data produced in the present work are contained in the manuscript

http://maps.org/mapp1

## Acknowledgements

The authors would like to thank Stephanie Kimble for her help in organizing this manuscript; and MPBC staff members Scott Hamilton, PhD (Director of Biostatistics) for statistical consultation; Glen Robinson (Clinical Data Assistant) and Chelsea Pamplin (TMF Associate) for conducting thorough quality checks on data entry in tables; Christina Faulk, MPH (Data Management Assistant) for creating figures; and Allison Coker, PhD for review of earlier manuscript drafts.

## Supporting Information

S1 Table. Mean (SD) Self-experience Scores among PTSD CAPS-5 Responders vs. Non-responders by Treatment Group.

